# Development of an electronic decision aid tool to facilitate mainstream genetic testing in ovarian cancer patients

**DOI:** 10.1101/2023.08.11.23293986

**Authors:** Kristen M. Shannon, Devanshi Patel, Jessica Jonas, Erica L. Blouch, Stephanie Hicks, Mackenzie Wooters, Meredith Seidel, Carly F. Grant, Margaret Emmet, Daniel C. Chung, Karen Sepucha

**Affiliations:** Massachusetts General Hospital Center for Cancer Risk Assessment, Boston, MA, USA; Division of Gastroenterology, Massachusetts General Hospital, Boston, Massachusetts, USA; Health Decision Sciences Center, Massachusetts General Hospital Center, Harvard Medical School, Boston, MA, USA

**Keywords:** genetic testing, ovarian cancer, decision aid, genetic counseling, shared decision-making

## Abstract

**Background:** Multigene panel testing is an important component of cancer treatment plans and risk assessment, but there are many different panel options and choosing the most appropriate panel can be challenging for healthcare providers and patients. Electronic tools have been proposed to help patients make informed decisions about which gene panel to choose by considering their preferences and priorities.

**Materials and Methods:** An electronic decision aid tool was developed in line with the International Patient Decision Aids Standards (IPDAS) collaboration. The multidisciplinary project team collaborated with an external healthcare communications agency and the MGH Cancer Center patient family advisory committee (PFAC) to develop the decision aid. Surveys of genetic counselors and patients were used to scope the content, and alpha testing was used to refine the design and content.

**Results:** Surveys of genetic counselors (n=12) and patients (n=228) identified common themes in discussing panel size and strategies for helping patients decide between panels and in identifying confusing terms for patients and distribution of patients’ choices. The decision aid, organized into two major sections, provides educational text, graphics and videos to guide patients through the decision-making process. Alpha testing feedback from the PFAC (n=4), genetic counselors (n=3) and a group of lay people (n=8) identified areas to improve navigation, simplify wording and improve layout.

**Conclusion:** The decision aid developed in this study has the potential to facilitate informed decision-making by patients regarding cancer genetic testing. The distinctive feature of this decision aid is that it addresses the specific question of which multigene panel may be most suitable for the patient. Its acceptability and effectiveness will be evaluated in future studies.

**Implications for Practice:** The use of multigene panel testing in oncology has increased significantly, but selecting the most appropriate panel for each patient can be challenging for healthcare providers and patients. The complexity and heterogeneity of the data generated by these tests, coupled with the potential psychological, emotional, and financial impacts, make informed consent a critical component of the genetic testing process. With the expansion of germline genetic testing, the traditional model of pre-test informed consent by a genetic counselor may no longer be scalable. Electronic decision aids have shown promise in increasing patient knowledge and empowerment in the shared decision-making process. The development of a comprehensive electronic tool can facilitate patient education while maximizing patient autonomy, allowing for more personalized and informed decisions about which gene panel to choose.⍰

## Introduction

Germline genetic testing has become a critical component of oncology treatment plans for many patients, including those with breast, ovarian, colon, pancreas, and other solid tumors. One of the most widely used types of genetic testing is multigene panel testing, which can simultaneously analyze multiple genes associated with a particular disease or a group of related conditions. While multigene panel testing can provide important diagnostic and therapeutic information, choosing the most appropriate panel for each patient can be a challenging task for healthcare providers and patients. This is because different tests may cover different sets of genes, have varying sensitivity and specificity, and offer various levels of clinical utility, among other factors. As a result, the selection of the optimal test can be a complex and daunting task, which requires careful consideration of multiple clinical and genetic factors.

Multigene panel testing can generate a large amount of complex and heterogeneous data, which can be difficult for healthcare providers and patients to interpret and utilize effectively. As highlighted by a recent review by Leenen et al. (2021), the interpretation and communication of genetic test results can be particularly challenging for patients, who may struggle to understand the implications of the results for their health and the health of their relatives. In addition, the choice of a specific multigene panel test can have important implications for patients, including psychological and emotional impacts, financial costs, and potential risks and benefits associated with the test. The variety of multi-gene cancer panel options complicates the process of informed consent as it relates to patient autonomy in medical decision-making (del Carmen, 2005). This critical element of the genetic testing process allows the patient to decide which, if any, multi-gene panel they would like to pursue, as aligned with their values. Genetic counselors typically invest a significant amount of time facilitating the detailed educational component and decision-making process for each patient considering multigene panel testing.

The clinical indications for germline genetic testing are expanding at a rapid pace, and germline genetic testing for all patients with solid tumors may soon be a reality (Liu, 2021). The traditional model of pre-test informed consent performed by a genetic counselor is not scalable given the limitations in the genetic counseling workforce (Esplin, 2022). Mainstream genetic testing is becoming an accepted alternative model, in which patients undergo pre-test genetic counseling and informed consent by their non-genetics healthcare provider (Bokkers, 2022). According to a study by Best et al. (2018), however, healthcare providers reported they lacked the necessary knowledge and expertise to choose the most appropriate multigene panel test for their patients, and often relied on personal experience or expert opinions to make a decision.

Electronic patient-facing educational tools have been developed to facilitate genetic testing and have been shown to increase patient knowledge and empowerment surrounding decision-making (Cragun, 2020). Furthermore, studies show that clinicians using decision aids (DAs) report improved quality of care and satisfaction in the shared decision-making process (Sepucha, 2016). However, most tools address the decision to pursue one genetic testing option rather than which multigene panel to select among the many options. Given the promising preliminary data, we proposed the development of a comprehensive electronic tool for ovarian cancer patients to facilitate patient education while maximizing patient autonomy. Our goal was to have patients select one of four options: pursue a smaller panel of clinically actionable genes related to ovarian cancer, pursue a medium-sized panel of mostly clinically actionable genes across cancer types, pursue a large panel of both clinically actionable and non-actionable genes across cancer types, or decline genetic testing. Here we describe the design of this electronic decision aid tool designed to help patients to make more informed and personalized decisions about which gene panel to choose by considering their preferences, values, and priorities.

## Methods

### Rationale for Decision Aid

Massachusetts General Hospital (MGH) is a large academic medical center in Boston, Massachusetts, part of the Mass General Brigham healthcare system. As a comprehensive cancer center, MGH Cancer Center provides expert care for patients with diverse types of cancer, including ovarian cancer. Most patients with ovarian cancer are treated by highly specialized gynecologic oncologists. The MGH Center for Cancer Risk Assessment (CCRA) provides genetic counseling and testing services to a wide variety of patients with and without cancer. Despite a staff of 14 genetic counselors, the number of cancer patients requiring access to genetic testing was outpacing the clinical capacity. Thus, we explored the possibility of mainstreaming genetic testing in ovarian cancer patients, meaning the patient would be educated and consented for genetic testing by the treating gynecologic medical oncologist.

When we proposed mainstream genetic testing model to the gynecologic oncology providers at our institution, they identified two main concerns with the mainstream approach. They expressed concern for a lack of expertise in providing pre-test patient education and in helping patients choose the most appropriate gene panel. Their bigger concern, however, was the amount of additional clinic visit time required given the lengthy nature of the informed consent process. We sought to alleviate these two concerns by developing a patient-friendly intervention to assist in the patient education and decision-making process.

### Multi-step framework

Our decision aid was developed in line with the framework described by Coulter et al. and put forth by the International Patient Decision Aids Standards (IPDAS) collaboration, which consists of five steps: (1) defining the scope, (2) design, (3) prototype development, (4) alpha testing and steering committee review, and (5) beta testing [Coulter 2013]. Here we focus on the first four steps; beta testing is occurring in both a pilot study and as a randomized controlled trial and will be reported separately.

#### 1. Defining the Scope

Our decision aid focused on a target audience of ovarian cancer patients for three main reasons. First, germline genetic testing for ovarian cancer patients is universally recommended (NCCN 2022), obviating the need to determine eligibility for testing based on family history or other factors. Second, because of the universal recommendation, most insurance companies cover the cost of germline genetic testing for ovary patients, mitigating the discussion of out-of-pocket cost. Finally, ovarian cancer is a relatively uncommon diagnosis, with <100 new patients diagnosed at our institution each year, making this an ideal population in which to pilot a mainstream genetic testing model.

Our project team was comprised of genetic counselors, medical oncologists, decision scientist and health communication experts. As is consistent with our current clinical genetic counseling practice, the team wanted a DA (Decision Aid) that provided multiple options for genetic testing be offered to patients. Throughout, we wanted the DA to use a combination of written information, graphics, and audio/video content to accommodate various learning styles. We planned to include patient quotes as part of the educational strategy. Finally, following IPDAS, we wanted to include a values clarification exercise and a summary component to assist patients in decision-making regarding three panel options for genetic testing (Figure 1).

**Figure 1.**
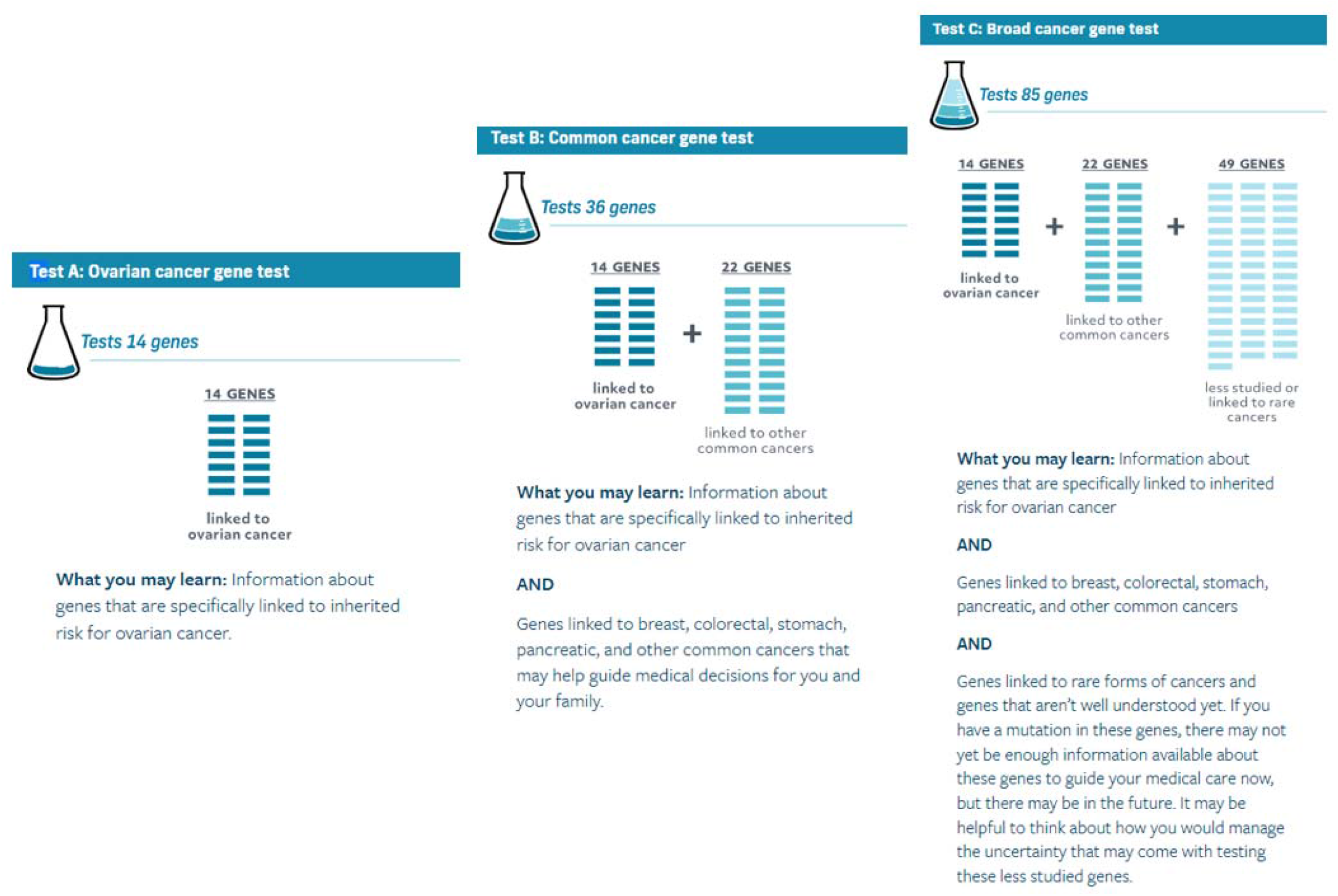
Gene Panel Testing Options

After an extensive literature review, we found there were various genetic testing DAs available, but few were truly interactive or designed for patient use (Williams et al 2008; Birch 2015). In addition, most DAs for cancer predisposition testing were tailored to syndrome-specific/gene-specific testing (Peterson et al 2004; Sherman et al 2018). Our team reached out to the National Society of Genetic Counselors (www.nsgc.org), who expressed support for this unique project. We secured an educational grant from Astra Zeneca (Grant ID# 41473887) for the development of the electronic DA.

#### 2. Design

Our project team collaborated with an external healthcare communications agency (Health Communication Core (HCC)) and the MGH Cancer Center Patient Family Advisory Committee (PFAC) to identify key information about the germline genetic testing and informed consent process that would be essential to patient understanding.

The HCC held discussions with the project team to clarify how the DA would be integrated into the clinical flow and underwent mock counseling sessions with the CCRA genetic counselor staff to determine how best to replicate in-person counseling in the decision aid. Once the major educational topics were identified, we sought to determine the appropriate ‘flow’ of the aid and decided to gather more information on current practice models. Our project team investigated how genetic counselors were facilitating patient decision-making in the traditional setting, and we conducted an online anonymous survey of the CCRA genetic counselors. Two terms were defined, and the 3 questions were asked in an open-ended text box (Figure 3).

Finally, the Project team developed an IRB (Institutional Review Board) study (MGB IRB#2019P000342) to survey patients and genetic counselors to learn more about how individuals make decisions about genetic testing. The study aimed to describe the frequency with which patients choose one of several panel options or to decline genetic testing, and to identify variables that may predict patient choice (Emmet et al, submitted). One goal of this survey project was to identify values-based assessments that could be used in the development of our DA.

#### 3. Prototype Development

The design steps above resulted in the development of interactive educational content and values-clarification sections that reviewed and reinforced information while encouraging users to make a decision. Patient responses were compiled into a summary patients could opt to have emailed to them before they exited the DA. Videos of genetic counselors explaining key concepts were produced by an outside vendor. Audio “testimonials” of patients discussing their decision-making process and representing diverse decisions were obtained. The HCC developed easy to understand graphics illustrating cancer risk and other genetic testing concepts. Users of the DA were able to move through the decision aid sequentially, access optional in-depth content, skip or return to specific sections, or jump to the “choose a genetic testing panel,” based on their personal preferences, level of knowledge, and readiness to decide. Users could also add questions and comments as they moved through the DA, which were then compiled in the downloadable summary.

#### 4. Alpha testing and project team review

To refine the prototype, we conducted alpha testing with PFAC members and genetic counselor project members. Each PFAC member met individually with a member of HCC who used a semi-structured interview guide to gather information on the readability, relevance, and acceptability of the decision aid as the PFAC member navigated through the tool. Each genetic counselor project member tested the decision aid and provided feedback on readability, relevance, and acceptability. In addition, we informally had 8 lay people navigate the decision aid and provide feedback on readability and acceptability. The iterative process of revising the DA was overseen by a multidisciplinary steering committee consisting of the project team and the HCC team.

## Results

### DA Design

The HCC and project team held a one-hour focus group with 3 (female) members of the PFAC. After an introduction to the project, the PFAC members were asked general questions about genetic testing, how they accessed information online, and their thoughts on how to explain complex scientific information to lay people (Figure 2). The focus group was audio recorded and transcribed. The PFAC contributions to the future DA development included ideas on readability, the ratio of text and video content, and key messages (e.g., impact on family).

**Figure 2.**
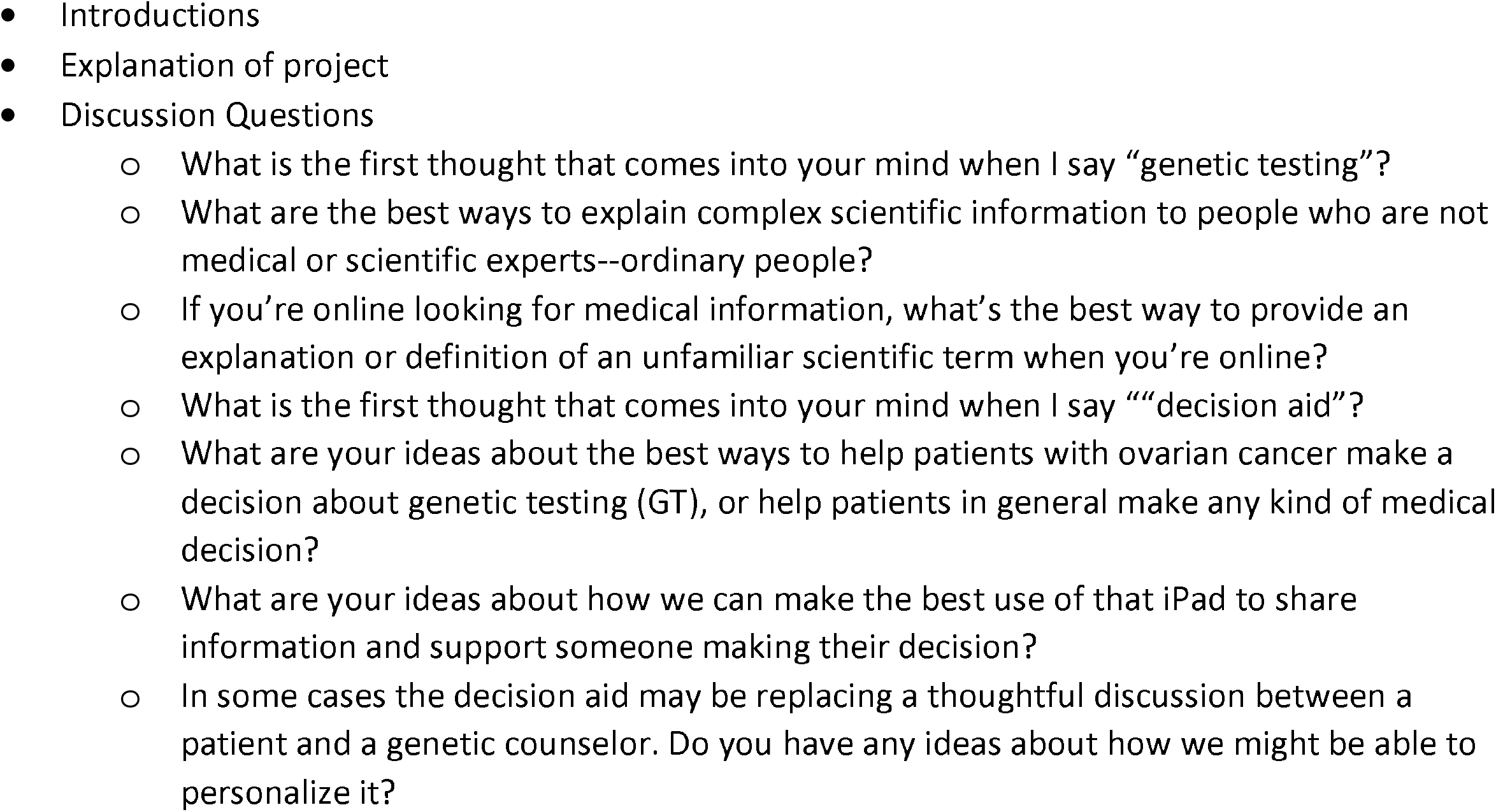
PFAC Focus Group Discussion Guide

**Figure 3.**
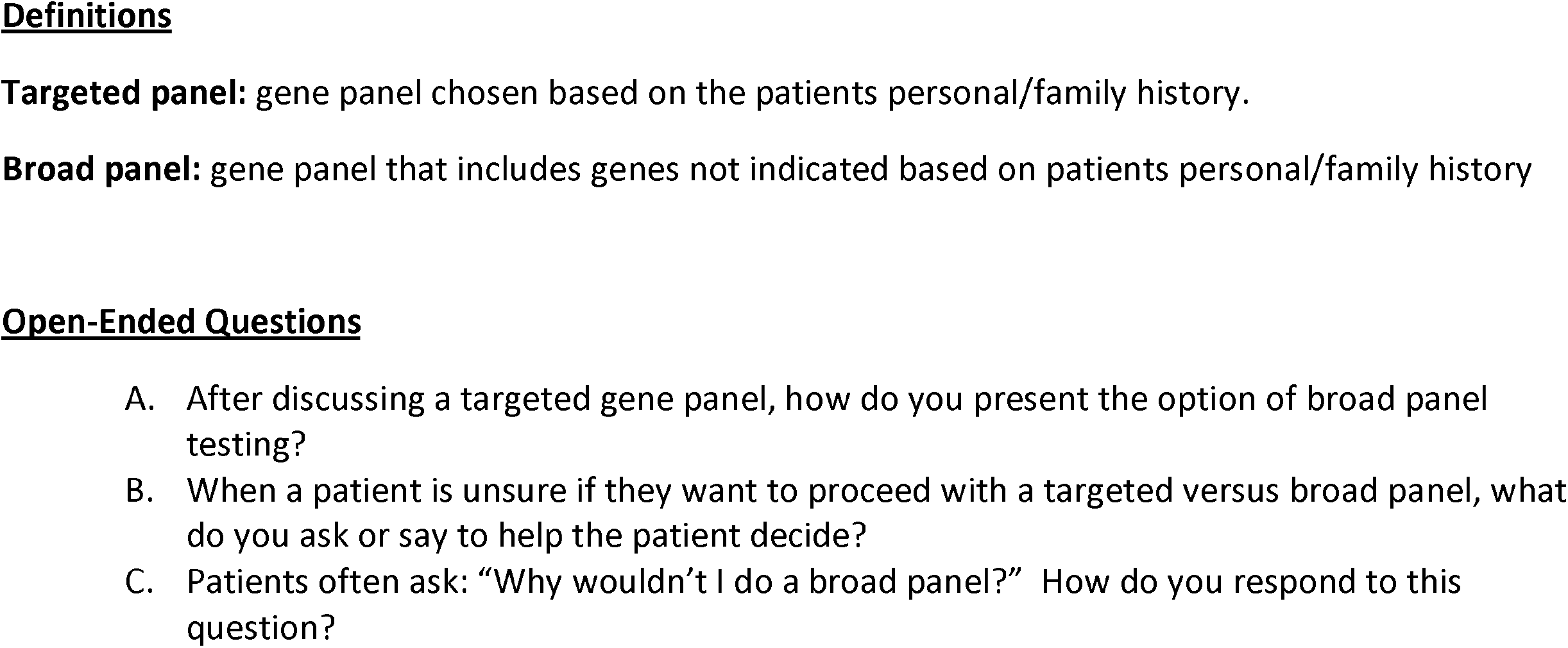
: Current practice of Genetic Counselor Survey

The key educational content was ascertained from the genetic counselor team members. The content underwent four revisions by three genetic counselors on the project team before being sent to HCC for plain language support. The topics can be seen in Figure 4 with brief descriptions of content type.

**Figure 4.**
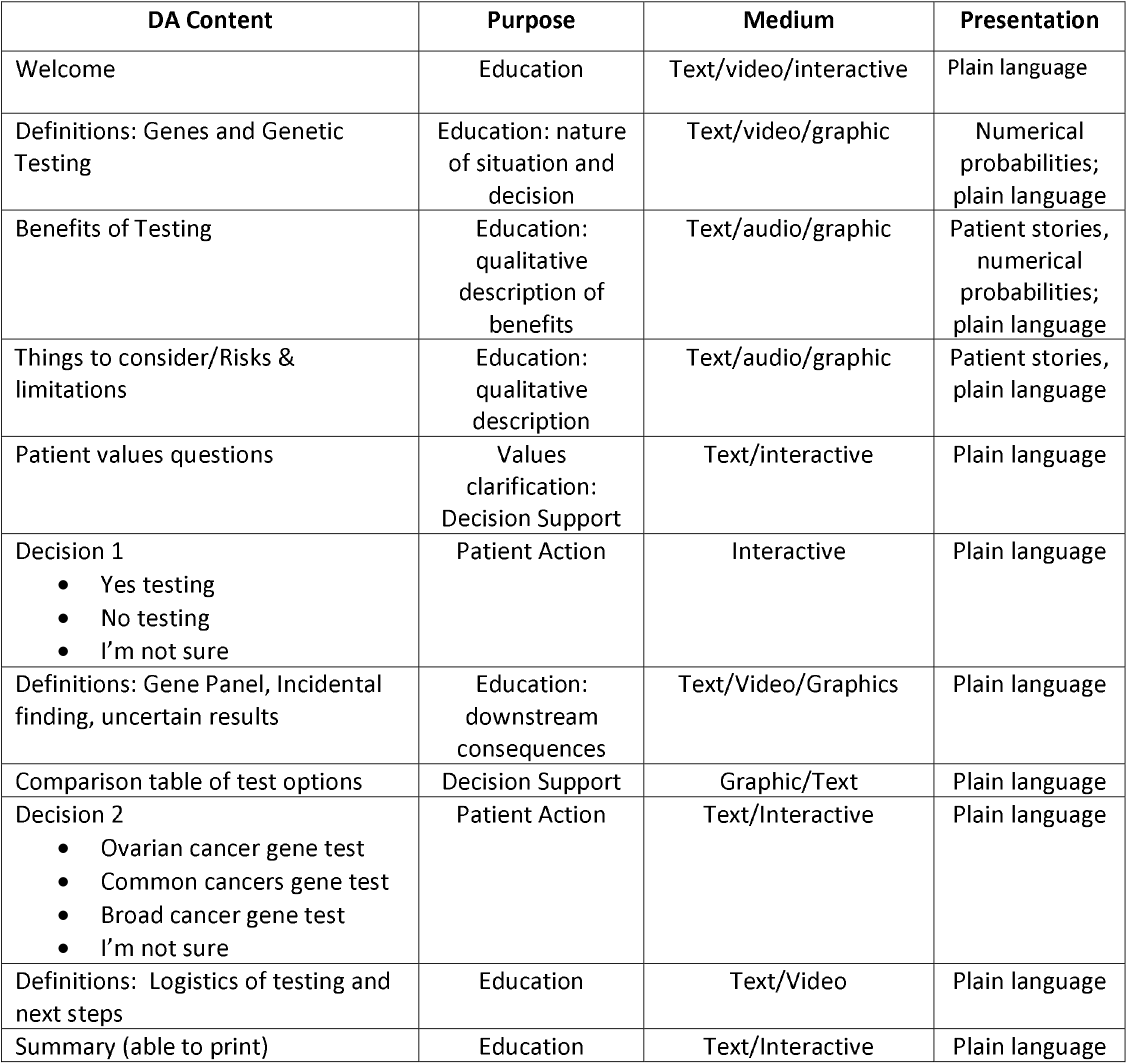
Electronic Decision Aid Content and Purpose of Content.

When surveying the genetic counselors regarding current practice issues, 12 CCRA genetic counselors responded to an anonymous survey. Common themes were identified and categorized. Most genetic counselors (10/12) started the discussion about panel size with a description of targeted genetic testing and then expanded to include broad genetic testing panel options. One genetic counselor reported discussing the broadest panel first and then ‘worked backwards’ to the targeted panel, and one genetic counselor discussed a broad panel as a reflex option in the context of a negative targeted panel. When asked for strategies for facilitating patient decision-making, genetic counselors most commonly explored how the patient dealt with uncertainty (8/12); how the patient felt about getting an unexpected but clinically relevant finding (5/12) and if the patient would describe themselves as an ‘information seeker’ (4/12). Other strategies included exploring if the patient found medical information particularly anxiety provoking, if they were someone who needed to know the ‘latest and greatest’, if they could compartmentalize information well, and if they were comfortable with information changing over time. Finally, when asked ‘why wouldn’t I do a broad panel?’, genetic counselors most often cited three patient perspectives: 1. Some people didn’t like uncertainty (6/12); 2. Some patients were concerned about a higher rate of a variant of uncertain significance (VUS) result in the broad panel (6/12); and 3. Some patients only wanted information relevant to their current circumstances (5/12). These genetic counselor survey responses informed the educational components in the DA (fig 2) and in some of the text used to describe the risks and benefits of different panel types.

Our patient survey study (Partners Healthcare IRB Protocol #2019P000342) was designed to explore patient choice between a full spectrum of multi-gene panel options (Emmet et al, submitted). It examined factors associated with the selection of a specific multi-gene panel test by patients when undergoing pre-test education and informed consent with a genetic counselor. The study confirmed the importance of patient choice, provided insight into the distribution of patients’ preferences for testing, and supported the idea that a decision aid may be helpful to facilitate informed decision-making.

### Prototype

The full tool is available at https://mghcancergeneticsda.com/. The project team organized the information into two major sections: 1) deciding about genetic testing (yes/no) and 2) deciding which panel type (ovarian cancer, common cancer, broad cancer). Each major heading is followed by educational points that expand in an ‘accordion’ style to provide additional context. Provider videos support the learning process and patient quotes are used to provide insight into the patient experience with genetic testing.

The learning process focuses first on the decision to have genetic testing (versus not having genetic testing) and includes value statements that the user rates on a 7-point Likert scale. The user is then prompted to decide if they would like to proceed with genetic testing. If the user decides they do not want genetic testing, they can either end the DA tool or continue to learn more about testing options. The user can also indicate they are not ready to decide about genetic testing. If this is the case, they are asked if they would like to speak with their oncologist, speak with a genetic counselor, or take some time to think about it. Finally, if the user decides to proceed with testing, they are guided through the second educational aspect of the DA: education and decision-making regarding which gene panel to pursue. Users are provided with the three testing options, including the limitations and benefits of each panel type (Figure 1). Once the education is completed, the patient is prompted to choose one of three tests. The final education section of the tool provides testing logistics and next steps (e.g., getting a blood draw, turnaround time of test). Once the DA is completed (whether a test is chosen), the user receives summative guidance about which decision they have made and can email it to themselves for their records.

### Alpha testing

Four members of the PFAC assisted with the Alpha testing. Three of these (female) individuals had participated in the design focus group and one (male) was a new member. PFAC members suggested changes-based acceptability and readability. Changes to the the decision aid because of the PFAC testing included adding of a ‘welcome screen’ at the outset of the DA, moving the navigation bar to the top of the screen and the creating additional ‘accordion’ buttons to reduce the amount of text on a page. Changes to readability included removing redundant text and labeling statistical information more clearly.

The eight lay people who navigated the decision aid and all agreed the content was relevant and readable, but suggested text modifications to reduce redundancy. Modifications were made to the navigation based on their feedback, including the addition of a ‘back’ button to return to a previous page or section, and a re-design of the ‘accordion’ feature to make it more readable.

The genetic counselor project team members identified minor navigation concerns (e.g., when clicking a button, it did not skip to the correct location) which were rectified with programming.

## Discussion

We describe the development of a decision aid for multigene panel cancer genetic testing that facilitates informed decision-making by patients. The distinctive feature of this decision aid is its ability to tackle the intricate task of facilitating selection of the most suitable multigene panel test, a challenge that has been widely recognized in the medical community. As noted in a recent review by Tung et al. (2021), the increasing number of available multigene panel tests has created a significant dilemma for clinicians, who struggle to identify the most appropriate option for each patient’s unique clinical scenario. This decision aid tool offers a comprehensive and personalized approach to guide clinicians towards a more informed and evidence-based decision-making process.

The educational content of the decision aid underwent several revisions, and patient surveys were conducted to explore factors associated with the selection of a specific multigene panel test. A patient survey study performed during the development process showed that while large panels were common, testing choice was distributed across different panel options and was difficult to predict (Emmet et al., submitted). This highlights the importance of patient choice and the need for a decision aid to facilitate informed decision-making.

The alpha testing phase included feedback from patient and family advisory council (PFAC) members and genetic counselor project team members. PFAC members provided significant feedback resulting in major navigation and content changes. Minor navigation concerns were identified by the genetic counselors, and rectified, and lay people navigated the decision aid and suggested no changes. This feedback indicates that the decision aid is user-friendly and can be used by a wide range of patients.

The tool has certain limitations that should be noted. First, decision aids may not be appropriate for all patients, particularly those with complex medical histories or co-morbidities (Finch et al., 2021). Further, our decision aid is currently available in only one language and designed for a specific disease population, namely ovarian cancer. It is worth noting that plans are in place to address these limitations. Consistent with research by Yu et al. (2020) which highlights the importance of considering cultural and linguistic factors when developing decision aids for diverse patient populations, we will be translating the tool into multiple languages. In addition, we will be broadening its applicability to other cancer types.

In conclusion, the decision aid developed in this study has the potential to facilitate informed decision-making by patients regarding cancer genetic testing. The decision aid’s effectiveness and impact on patient decision-making will be evaluated in future studies to validate its utility.

## Data Availability

All data produced in the present study are available upon reasonable request to the authors

## Acknowledgements

We thank Linda Rodgers-Fouche, MGc, CGC who assisted with the development of the gene panels referred to in the DA.

We thank Dana-Farber/Harvard Cancer Center in Boston, MA, for the use of the Health Communication Core, which provided content development and technical development service. Dana-Farber/Harvard Cancer Center is supported in part by an NCI Cancer Center Support Grant # NIH 5 P30 CA06516

## Supplementary Information

n/a

Contact and competing interest information for all authors

Kristen M. Shannon, MS, CGC

Mass General Cancer Center

Center for Cancer Risk Assessment

55 Fruit St. – YAW 10B

Boston, MA 02114

617-726-9337 (ph)

617-726-9418 (fax)

No competing interests

Devanshi Patel, MS, CGC

Mass General Cancer Center

Center for Cancer Risk Assessment

55 Fruit St. – YAW 10B

Boston, MA 02114

617-724-3285 (ph)

617-726-9418 (fax)

No competing interests

Jessica Jonas, MS, CGC

Mass General Cancer Center

Center for Cancer Risk Assessment

55 Fruit St. – YAW 10B

Boston, MA 02114

617-724-1971 (ph)

617-726-9418 (fax)

No competing interests

Erica L. Blouch, MS, CGC Mass General Cancer Center

Center for Cancer Risk Assessment

55 Fruit St. – YAW 10B

Boston, MA 02114

617-724-1971 (ph)

617-726-9418 (fax)

No competing interests

Stephanie Hicks, MS, CGC

Mass General Cancer Center

Center for Cancer Risk Assessment

55 Fruit Street YAW 10B

Boston, MA 02114

617-724-5538

617-726-9418 (fax)

No competing interests

Mackenzie Wooters, MS

Mass General Cancer Center

Center for Cancer Risk Assessment

55 Fruit Street YAW 10B

Boston, MA 02114

617-726-9323

617-726-9418 (fax)

No competing interests

Meredith Seidel, MS, CGC

Mass General Cancer Center

Center for Cancer Risk Assessment

55 Fruit Street YAW 10B

Boston, MA 02114

617-724-0110

617-726-9418 (fax)

No competing interests

Carly F. Grant, MS, CGC

Mass General Cancer Center

Center for Cancer Risk Assessment

55 Fruit Street YAW 10B

Boston, MA 02114

617-724-1971

617-726-9418 (fax)

No competing interests

Margaret Emmet, MS, CGC

Mass General Cancer Center

Center for Cancer Risk Assessment

55 Fruit Street YAW 10B

Boston, MA 02114

617-724-1971

617-726-9418 (fax)

No competing interests

Daniel C. Chung, MD

Gastroenterology Division

Center for Cancer Risk Assessment

55 Fruit Street

Boston, MA 02114

617-726-8687

No competing interests

Karen Sepucha, PhD

Health Decision Sciences Center, 100C-16

55 Fruit Street

Boston, MA 02114

617 724-3350

No competing interests

Data sharing plans (including all data, documentation, and code used in analysis)

n/a

## Funding information

Funding for the development of the electronic tool described here was generously obtained by Astra Zeneca (Educational Grant ID# 41473887).

The authors did not receive any compensation from Astra Zeneca, all funds were directed to the outside contractors for the development of the tool.

- Introductions
- Explanation of project
- Discussion Questions
  - What is the first thought that comes into your mind when I say “genetic testing”?
  - What are the best ways to explain complex scientific information to people who are not medical or scientific experts--ordinary people?
  - If you’re online looking for medical information, what’s the best way to provide an explanation or definition of an unfamiliar scientific term when you’re online?
  - What is the first thought that comes into your mind when I say ““decision aid”?
  - What are your ideas about the best ways to help patients with ovarian cancer make a decision about genetic testing (GT), or help patients in general make any kind of medical decision?
  - What are your ideas about how we can make the best use of that iPad to share information and support someone making their decision?
  - In some cases the decision aid may be replacing a thoughtful discussion between a patient and a genetic counselor. Do you have any ideas about how we might be able to personalize it?

## Definitions

Targeted panel: gene panel chosen based on the patients personal/family history.

Broad panel: gene panel that includes genes not indicated based on patients personal/family history

### Open-Ended Questions

A. After discussing a targeted gene panel, how do you present the option of broad panel testing?
B. When a patient is unsure if they want to proceed with a targeted versus broad panel, what do you ask or say to help the patient decide?
C. Patients often ask: “Why wouldn’t I do a broad panel?” How do you respond to this question?

## References

Best, D. H., O’Neill, S. C., & Lyon, E. (2018). Genomic medicine in the era of multigene panels: successes, complexities, and challenges. Advances in Clinical Chemistry, 83, 1–32. doi: 10.1016/bs.acc.2018.03.001

Birch, P. H. (2015). The state of shared decision-making tools for high-risk medical decisions. JAMA Internal Medicine, 175(12), 2013–2014. doi: 10.1001/jamainternmed.2015.5490.

Bokkers K, Vlaming M, Engelhardt EG, et al. The Feasibility of Implementing Mainstream Germline Genetic Testing in Routine Cancer Care-A Systematic Review. Cancers (Basel). 2022;14(4):1059. Published 2022 Feb 19. doi:10.3390/cancers14041059

Coulter, A., Stilwell, D., Kryworuchko, J., Mullen, P. D., & Ng, C. J. (2013). A systematic development process for patient decision aids. BMC Medical Informatics and Decision Making, 13(Suppl 2), S2. doi: 10.1186/1472-6947-13-S2-S2

Cragun D, Weidner A, Tezak A, Zuniga B, Wiesner GL, Pal T. A Web-Based Tool to Automate Portions of Pretest Genetic Counseling for Inherited Cancer. J Natl Compr Canc Netw. 2020;18(7):841–847. doi:10.6004/jnccn.2020.7546

del Carmen MG, Joffe S. Informed consent for medical treatment and research: a review. Oncologist. 2005;10(8):636–641. doi:10.1634/theoncologist.10-8-636

Emmet, M., Chang, Y., Chung, D.C., Caruso, A.R., Shannon, K.M. (submitted) Patient Decisions Regarding Cancer Gene Panel Testing: An Exploratory Study. Journal of Genetic Counseling.

Esplin ED, Nielsen SM, Bristow SL, et al. Universal Germline Genetic Testing for Hereditary Cancer Syndromes in Patients With Solid Tumor Cancer. JCO Precis Oncol. 2022;6:e2100516. doi:10.1200/PO.21.00516

Finch T, Rapley T, Girling M, et al. Improving the quality of decision-making in complex surgery: A qualitative study exploring the attitudes and experiences of patients and clinicians to using a patient decision aid. PLoS One. 2021; 16(1): e0243743. doi:10.1371/journal.pone.0243743

Leenen, J., Henneman, L., & Timmermans, D. R. (2021). Genetic testing: communicating risks and results to patients. Expert Review of Molecular Diagnostics, 21(2), 199–206. doi: 10.1080/14737159.2021.1875461

Liu YL, Stadler ZK. The Future of Parallel Tumor and Germline Genetic Testing: Is There a Role for All Patients With Cancer?. J Natl Compr Canc Netw. 2021;19(7):871-878. Published 2021 Jul 28. doi:10.6004/jnccn.2021.7044

National Comprehensive Cancer Network (NCCN). Genetic/Familial High-Risk Assessment: Breast, Ovarian, and Pancreatic. (Version 1.2023). https://www.nccn.org/guidelines/guidelines-detail?category=2&id=1503. Accessed 12/21/22.

Peterson, E. A., Milliron, K. J., Lewis, K. E., Goold, S. D., Merajver, S. D., & Zauber, A. G. (2004). Communication outcomes of BRCA1/2 genetic counseling/testing in young breast cancer survivors. Cancer Epidemiology, Biomarkers & Prevention, 13(5), 867–873.

Sepucha KR, Simmons LH, Barry MJ, Edgman-Levitan S, Licurse AM, Chaguturu SK. Ten Years, Forty Decision Aids, And Thousands Of Patient Uses: Shared Decision Making At Massachusetts General Hospital. Health Aff (Millwood). 2016;35(4):630–636. doi:10.1377/hlthaff.2015.1376

Sherman, K. A., Shaw, L. K., Winch, C. J., Harcourt, D., Boyages, J., Cameron, L. D., & Brown, P. (2018). Reducing decisional conflict and enhancing satisfaction with information among women considering breast reconstruction following mastectomy: results from the BRECONDA randomized controlled trial. Plastic and Reconstructive Surgery, 142(3), 537–550. doi: 10.1097/PRS.0000000000004620

Tung, N., Domchek, S. M., Stadler, Z., Nathanson, K. L., & Couch, F. J. (2021). Increasing the value of genetic testing in oncology practice: Recommendations from the Personalized Medicine Working Group of the All of Us Research Program. Cancer Epidemiology, Biomarkers & Prevention, 30(3), 423–431. doi: 10.1158/1055-9965.EPI-20-1102

Williams, J. K., Katapodi, M. C., Starkweather, A., Badzek, L., & Stubbs, C. (2008). Patient, provider, and system factors impacting decision-making in genetic counseling and risk assessment for hereditary breast and ovarian cancer: a scoping review. Clinical Genetics, 74(4), 255–274. doi: 10.1111/j.1399-0004.2008.01047.x

Yu, J., Yoo, H. J., & Kim, J. H. (2020). Development and validation of a decision aid for lung cancer screening based on patients’ decision-making preferences. BMC Medical Informatics and Decision Making, 20(1), 1–12. https://doi.org/10.1186/s12911-020-1054-4

